# Comparison of Prompt Engineering and Fine-Tuning Strategies in Large Language Models in the Classification of Clinical Notes

**DOI:** 10.1101/2024.02.07.24302444

**Authors:** Xiaodan Zhang, Nabasmita Talukdar, Sandeep Vemulapalli, Sumyeong Ahn, Jiankun Wang, Han Meng, Sardar Mehtab Bin Murtaza, Dmitry Leshchiner, Aakash Ajay Dave, Dimitri F. Joseph, Martin Witteveen-Lane, Dave Chesla, Jiayu Zhou, Bin Chen

## Abstract

The emerging large language models (LLMs) are actively evaluated in various fields including healthcare. Most studies have focused on established benchmarks and standard parameters; however, the variation and impact of prompt engineering and fine-tuning strategies have not been fully explored. This study benchmarks GPT-3.5 Turbo, GPT-4, and Llama-7B against BERT models and medical fellows’ annotations in identifying patients with metastatic cancer from discharge summaries. Results revealed that clear, concise prompts incorporating reasoning steps significantly enhanced performance. GPT-4 exhibited superior performance among all models. Notably, one-shot learning and fine-tuning provided no incremental benefit. The model’s accuracy sustained even when keywords for metastatic cancer were removed or when half of the input tokens were randomly discarded. These findings underscore GPT-4’s potential to substitute specialized models, such as PubMedBERT, through strategic prompt engineering, and suggest opportunities to improve open-source models, which are better suited to use in clinical settings.

## Introduction

Large Language Models (LLMs) are transforming many fields^1-2^. Built on the Transformer architecture^3^, these models employ billions, sometimes, trillions, of tokens and parameters, enabling them to accomplish a range of tasks that were previously considered unattainable. From answering straightforward questions to handling complex reasoning tasks, their performance is often astonishing. Inspired by the scaling law^4^, which revealed a positive correlation between model performance and the number of parameters employed, tech companies are increasingly investing in these models using their proprietary training sets and massive computing power. Some models could be accessed through the API, such as OpenAI’s GPT-3^5^ (trained with 175 billion parameters and 300 billion tokens) or the software packages, including Meta’s Llama^6^ (trained with 65 billion parameters and 1.4 trillion tokens), and Google’s PaLM^7^ (trained with 540 billion parameters and 780 billion tokens). While some specifications, like the number of parameters, are publicly disclosed, many technical intricacies remain proprietary. Moreover, the task performance highly relies on skilled user prompting.

In the realm of biomedicine, LLM applications are burgeoning. While pretrained models like GatorTron^8^, trained on >80 billion words extracted from de-identified clinical text, showcased their capabilities across multiple clinical Natural Language Processing (NLP) tasks, most of the efforts remained on the fine-tuning of open-source LLM models. For example, Med-PaLM was a fine-tuned model from PaLM for biomedical research and performed considerably well but remained inferior to clinicians^9^. Compared to fine-tuned biomedical pretrained language models, LLMs like GPT-3.5 and GPT-4 showed promise in biomedical semantic similarity and reasoning tasks but are less effective in information extraction and classification^10^. Despite these efforts, few studies have systematically evaluated various prompting and fine-tuning strategies in published LLMs and compared with human performance.

Building on our prior work^11^, where we used early versions of language models like BERT^12^ for identifying metastatic cancer patients through clinical notes, we aim to further this research. Metastatic cancer is a leading cause of cancer-related deaths, thus identifying patients with metastatic cancer for early intervention is crucial for improving survival. In the Electronic Health Record (EHR) systems like Epic, metastatic cancer is often inadequately defined and labeled^11^, therefore, identifying those patients from clinical notes is intriguing. Training language models from scratch for clinical notes is resource-intensive, making pre-trained models followed by domain-specific fine-tuning a more viable alternative. In our previous study, we investigated several BERT variations such as BioBERT^13^, clinicalBERT^14^, and PubMedBERT^15^, and found that fine-tuning on PubMedBERT yielded the best results. However, compared to those LLMs trained with billions of parameters, the 110 million parameters in BERT are quite small. Moreover, although we could use tools to identify important features in the BERT models, compared to LLMs trained on large corpus, their reasoning capabilities remained limited.

Prompt engineering is an integral part to enhance model output quality, especially in complex tasks demanding causal reasoning^16^. In general, there are three different types of methods: zero-shot, one-shot and few-shot. Giving clear and specific instructions, describing the overall context, utilizing explicit constraints, and asking to play roles are some techniques that can be employed to achieve better results^17-19^. It is difficult to recommend a best practice, but with iterative testing and refinement, prompts can be significantly improved. Recent studies demonstrate that utilizing structured prompts can elevate LLM performance in medical diagnostics, where precision and interpretability are essential^20-22^.

Therefore, in this study, using metastatic cancer identification as a case study, we explored multiple recommended prompting strategies as well as common LLMs. We also evaluated the impact of various key parameters and compared their performance with that of domain experts. By fusing domain knowledge and chains of thought, we proposed an optimal prompt for in-context zero-shot learning and observed its favorable outcomes.

## Methods

### Dataset and Data Preprocessing

We used the approach previously presented in^,23^ to prepare the dataset from MIMIC-III^24^. More details can be found in **Appendix A**. As a result, we created a dataset that includes 1,873 discharge summaries with 178 patients with metastatic cancer. Each record consists of a discharge summary and a label for metastatic cancer. We conducted multiple pre-processing steps on the discharge summaries by following the same strategy in^11^. We employed a 7:2:1 ratio to split the data into training, validation, and testing sets.

### Manual Annotation

We engaged a panel of three medical fellows, each either possessing a medical degree or currently undergoing medical training, to manually annotate the test set. The fellows were provided with concise guidelines on how to identify instances of metastatic cancer based solely on the patient’s discharge summaries, without the aid of any supplementary tools. For this study, ‘*metastatic cancer’* was defined according to criteria specified in the previous work^23^: “*Cancers with very high or imminent mortality (pancreas, esophagus, stomach, cholangiocarcinoma, brain); mention of distant or multi-organ metastasis, where palliative care would be considered (prognosis < 6 months)*”. This definition guided the annotation process to ensure consistency and accuracy in the identification of relevant cases. A designated coordinator shared a screen with the annotators and advanced to the subsequent page only upon unanimous agreement among the fellows. To ensure anonymity, all annotations were recorded without identifiers. In total, the panel successfully annotated 188 discharge summaries in the test set within six hours. On average, each multi-page summary required approximately two minutes for thorough review. The analysis of the annotated data was conducted using Python.

### LLMs

We utilized various LLMs including OpenAI’s GPT models and Meta’s Llama, to classify the presence of metastatic cancer within discharge summaries. The setup and deployment of these models, alongside prompt engineering, one-shot, and fine-tuning strategies, were designed to ensure a rigorous evaluation protocol.

### OpenAI’s GPT models

The deployment of OpenAI’s GPT models, specifically GPT-3.5 Turbo and GPT-4, was executed on Microsoft Azure cloud computing service. An Azure subscription was obtained, and a dedicated Resource Group was established to manage essential resources such as compute instances and storage services. These models were then deployed within this Resource Group, ensuring an organized and efficient management of cloud resources. Access to the models was facilitated via the OpenAI Application Programming Interface (API), which required authentication using a unique API key. To adhere to OpenAI’s API usage policies, a robust rate limiting and request batching mechanism was implemented. The interaction with the API was encapsulated within a Python function designed to handle API requests with a retry mechanism. This function was tailored to efficiently manage both high-volume requests and API-specific errors, such as rate limit exceedances. It ensured reliable and efficient predictions retrieval from the GPT models.

In GPT models, the temperature parameter is used to control the randomness and creativity of the generated text in a generative language model. It adjusts the probabilities of predicted words in the model’s SoftMax output layer. A lower temperature sharpens these probabilities, making the word with the highest probability more likely to be chosen. As a result, the output becomes more conservative and stable. We have set the temperature to be 0.2 to get a more deterministic response. We have also tested for different token sizes to determine whether the output differs in any way. We have first tried one prompt with a variety of token sizes ranging from (500, 1000, 1500, 2000, 3000, 4000) and observed a variation in the F1 scores and other performance metrics. To illustrate the changes in F1 scores, we ultimately chose 1500 tokens and 3000 tokens to apply to all prompts.

### Meta Llama

Meta offers Llama as an open-source LLM. Llama comes in two main versions, with version 1 (V1) being released in February 2023, and version 2 (V2) in July of the same year. V1 offers model sizes of 7B, 13B, 33B, and 65B, and V2 provides sizes of 7B, 13B, and 70B. In this paper, fine-tuning was conducted using the 7B model from Llama V1 due to the hardware constraints of using larger models. These experiments were conducted with the research purpose consent provided by Meta and were carried out using the Huggingface Module.

### Prompt Engineering

Initially, we created a baseline prompt (prompt 0) based on the suggestion from ChatGPT and common knowledge. Following the concept of chains of thought, we refined these prompts by adding instructional steps. We also incorporated a universal prompt, as suggested in a recent study that introduced zero-shot reasoners, by simply adding “*Let’s think step by step*” before each answer^25^. In total, we explored six different prompts.

#Prompt 0

“Please act as a curator. Based on the input discharge summary, classify if the patient has metastatic cancer or not. Please provide a concise response as either ‘Yes’ or ‘No’. “

#Prompt 1

“Please act as a curator. Based on the input discharge summary, classify if the patient has metastatic cancer or not using the following steps.

Step 1: identify if this patient has cancer or not.

Step 2: if this patient has cancer, identify its staging, grade, and primary site.

Step 3: if this patient has metastatic cancer, identify its primary site, and metastatic.

Give a final decision ‘Yes’ or ‘No’ only.”

#Prompt 2

“This is a discharge summary for a patient who underwent diagnostic tests, please act as a healthcare professional and classify if the patients has metastatic cancer or not using following steps:

1. Identify if a patient has cancer or not.
2. If this patient has cancer, identify if it’s metastatic cancer.
3. Based on the information please provide a concise response as either ‘Yes’ or ‘No’.”

#Prompt 3

“This is a discharge summary for a patient who recently underwent diagnostic tests for suspected metastatic cancer. Please analyze the following information and provide a concise response as either ‘Yes’ or ‘No’ based on the presence of only metastatic cancer in the patient’s discharge summary.”

#Prompt 4

“This is a discharge summary for a patient. Please act as a healthcare professional and provide a response based on the summary using the following instructions:

1. Identify if a patient has metastatic cancer or not.
2. If there is clear evidence of metastatic cancer, respond ‘Yes’.
3. If there is no strong evidence of metastatic cancer or if the evidence is unclear, respond ‘No.’

Please provide a concise response as either ‘Yes’ or ‘No’. “

#Prompt 5

“Please act as a curator and analyze the following discharge summary, classify if the patient has metastatic cancer or not. Let’s think step by step. Choose the final answer from the list ‘Yes’ or ‘No’.”

### Context in Zero-Shot, One-Shot Learning, and Fine-Tuning

To use LLMs for our target classification task, there are three approaches we study in this work. The first approach is zero-shot learning, which inputs a combination of our designed prompts and clinical notes on untouched LLMs without giving any example. We can also conduct one-shot learning, which adds a few examples of clinical note and prediction labels, in addition to the inputs used by zero-shot learning, again on untouched LLMs. Finally, we can revise LLMs by fine-tuning clinical notes from the training set and adjusting the distribution of LLMs through back-propagation.

We tested the proposed six prompts in a zero-shot learning setting. We found that with the proper construction of prompts, one shot learning did not offer any additional benefits, so we focused primarily on zero-shot learning. For the fine-tuning process, we utilized the Llama architecture. The Llama model, known for its versatility and efficiency, offers various sizes ranging from 7 billion to 65 billion parameters. In our research, we leverage the Llama-7B model, which has been adapted for compatibility with the Transformers/HuggingFace framework. To make it better suited for specific tasks, such as clinical note classification and other medical-related challenges, we contemplate further fine-tuning of the Llama-7B model. Technically, we further employ approaches such as Parameter-Efficient Fine-Tuning (PEFT)^26^, 4-bit floating-point quantization^27^, and low-rank adaptation (LoRA)^28^ in our fine-tuning process to make it more efficient. The number of fine-tuning epochs in our experiment is setting to be 3. And the batch size is 8. Within the optimization phase, we have chosen the AdamW(32-bit) optimizer, a widely adopted selection for fine-tuning large language models. Furthermore, for our experiments, we have standardized the token size within the Llama model to 2048 tokens, which is appropriate in our computing environment. The Llama fine-tuning experiment was executed within a high-performance computing environment, featuring four NVIDIA A5000 graphics cards, each having 24 gigabytes (GB) of GPU memory. This robust hardware configuration provided us with ample computational power and memory capacity to efficiently fine-tune the Llama-7B model on our training dataset.

### Evaluation

To assess the classification performance of the GPT models, we executed a rigorous evaluation protocol across various prompts and input sizes (1500 and 3000 tokens). This analysis included the utilization of both GPT-3.5 Turbo and GPT-4 models. For each prompt configuration, we performed five separate runs to ensure result consistency and reliability. The evaluations were done using input lengths of 1500 and 3000 tokens to determine the impact of input size on the classification quality. Key metrics such as F1 scores, recall, and precision were recorded for each run. The averages of these metrics were then computed and summarized in a comprehensive table, facilitating direct comparisons of the effectiveness of each prompt and input size on the model’s classification accuracy. For manual annotation, the inter-annotator agreement was measured using Fleiss’ Kappa, a statistical measure that accounts for agreement occurring by chance. The performance of each medical fellow annotator was individually assessed in terms of F1 score, recall, and precision to gauge the reliability of manual annotations in identifying metastatic cancer within the discharge summaries. Comparative analysis was conducted across all models and methods employed in the study, including PubMedBERT, and Meta Llama-7B. For each model and approach, we reported the F1 scores, recall, and precision metrics derived from the test set. This enabled a thorough comparative evaluation, highlighting the relative strengths and weaknesses of each method in the task of metastatic cancer classification from clinical notes. All the analyses were conducted using Python, leveraging libraries such as Pandas for data manipulation, Scikit-learn for model evaluation, and Huggingface’s Transformers for LLM interactions. This ensured a consistent and reproducible analysis environment.

## Results

### Data Statistics and Baseline Performance

Before training the model, the dataset was preprocessed and tokenized. The token counts for each row in the training, validation, and test sets were calculated to understand the distribution of the data and to ensure that it aligns with the model’s limitations. Within the training set, token counts ranged from a minimum of 5 tokens to a maximum of 5348 tokens. On average, each record contained approximately 1791 tokens, with a standard deviation of 1008 tokens. In the validation set, the token values spanned from a minimum of 27 to a maximum of 5401 tokens. The average token count for this set were around 1747 tokens, accompanied by a standard deviation of approximately 1004 tokens. Similarly, the test dataset displayed token counts that varied from a minimum of 15 to a maximum of 5104 tokens. On average, each row consisted of approximately 1824 length of tokens, and the standard deviation was approximately 973 tokens.

The PubMedBERT model was implemented utilizing the MetBERT code repository from GitHub^11^. In the identification of cases with metastasis, the precision was 0.790 and a recall of 0.610, resulting in an F1-score of 0.690. These metrics indicate the model’s effectiveness in correctly classifying instances with and without metastasis.

### Manual Annotation

In evaluating the reliability of manual annotations for metastatic cancer identification within discharge summaries, we examined inter-annotator agreement and individual annotator performance (**Table 1**). Using the percent agreement metric, we found an impressive 94% agreement among the three medical fellows, demonstrating the robustness of the manual annotation process and dataset validity. The high percent agreement suggests a shared understanding of annotation guidelines. Fleiss’ Kappa κ, indicating inter-rater agreement beyond chance, yielded a substantial value of 0.868, further confirming annotation reliability. Together, these metrics suggest that the annotators were highly competent in identifying metastatic cancer cases from patients’ discharge summaries.

**Table 1:**
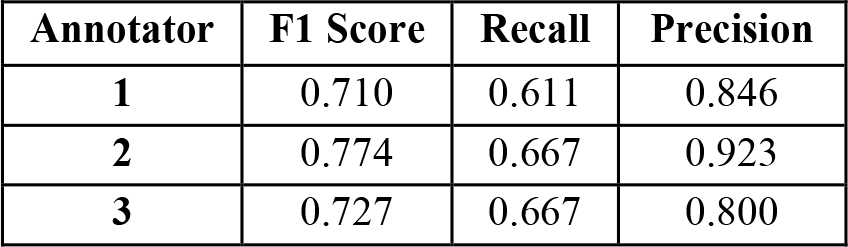
Evaluation Metrics of Annotator Performance Compared to Actual Cases.

### Zero-shot and One-shot Learning

We explored the GPT-3.5 Turbo and GPT-4 models using the test set. Regardless of the input token size and prompts, GPT-4 consistently outperformed GPT-3.5 Turbo, often by a large margin (**Table 2**).

**Table 2:**
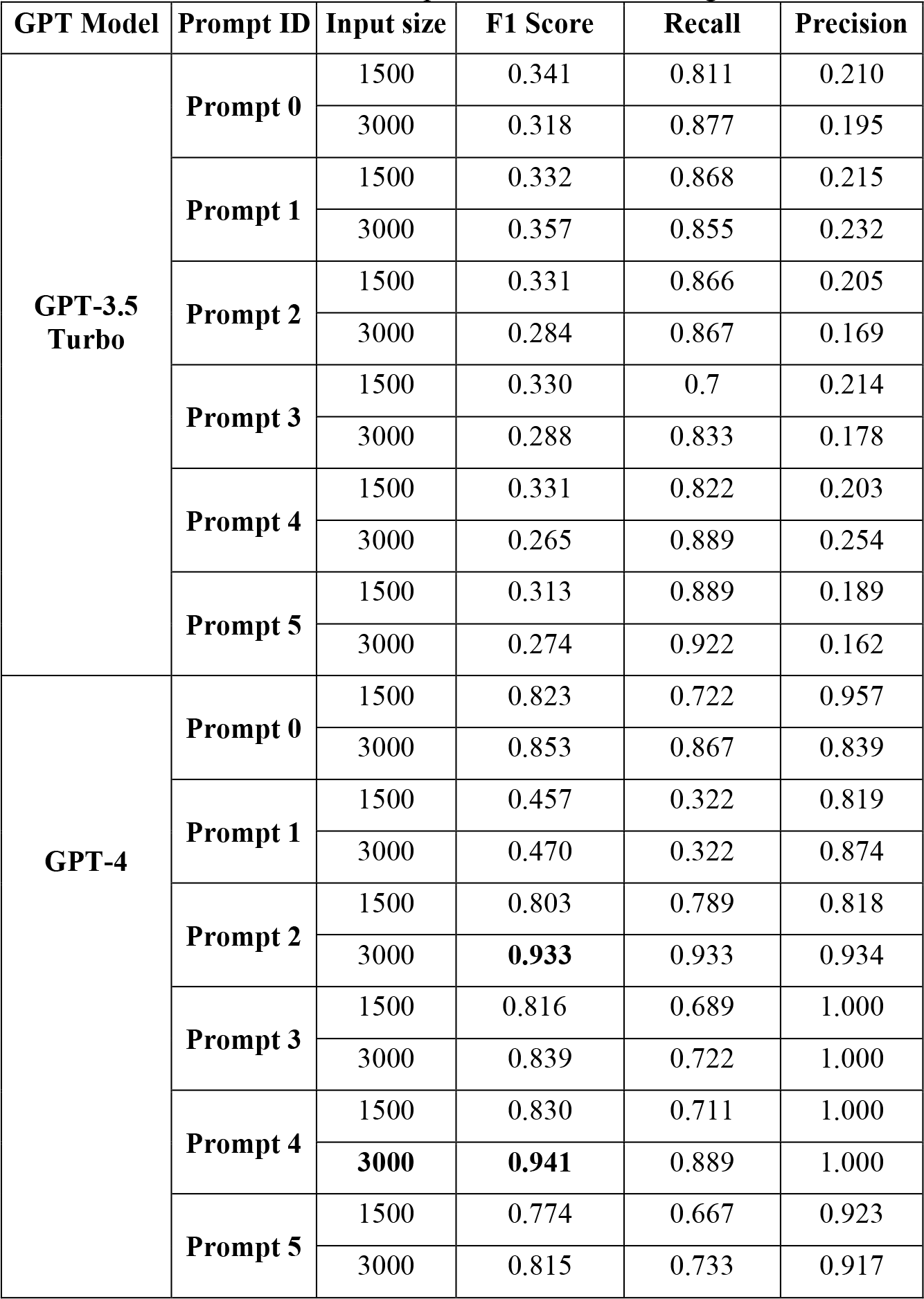
Evaluation of the Six Prompts in GPT Models Using the Test Set.

Among the six prompts, for both the token size prompt 4 yielded better results with an F1 score of 0.941 followed by prompt 2 with an F1 score of 0.933 in GPT-4. It is clear that prompts with structured instructions generally perform better, moreover simple prompts written in clear and concise way without any complexity also gives comparable performance. Notably, smaller tokens tend to result in better performance than longer tokens in GPT-3.5 Turbo. We observed that GPT-4 handles longer input sizes (3000 tokens) effectively, often outperforming its handling of shorter input sizes (1500 tokens). This is particularly evident with prompt 4, where GPT-4 achieved an F1 Score of 0.941 and precision of 1.000 at an input size of 3000, suggesting a remarkable ability to maintain high precision with increased input length.

**Figure 1** showed an example of the discharge summaries of a metastatic cancer patient along with the reasoning from GPT. GPT-4 correctly identified metastatic cancer based on the presence of key factors in the discharge summaries.

**Figure 1:**
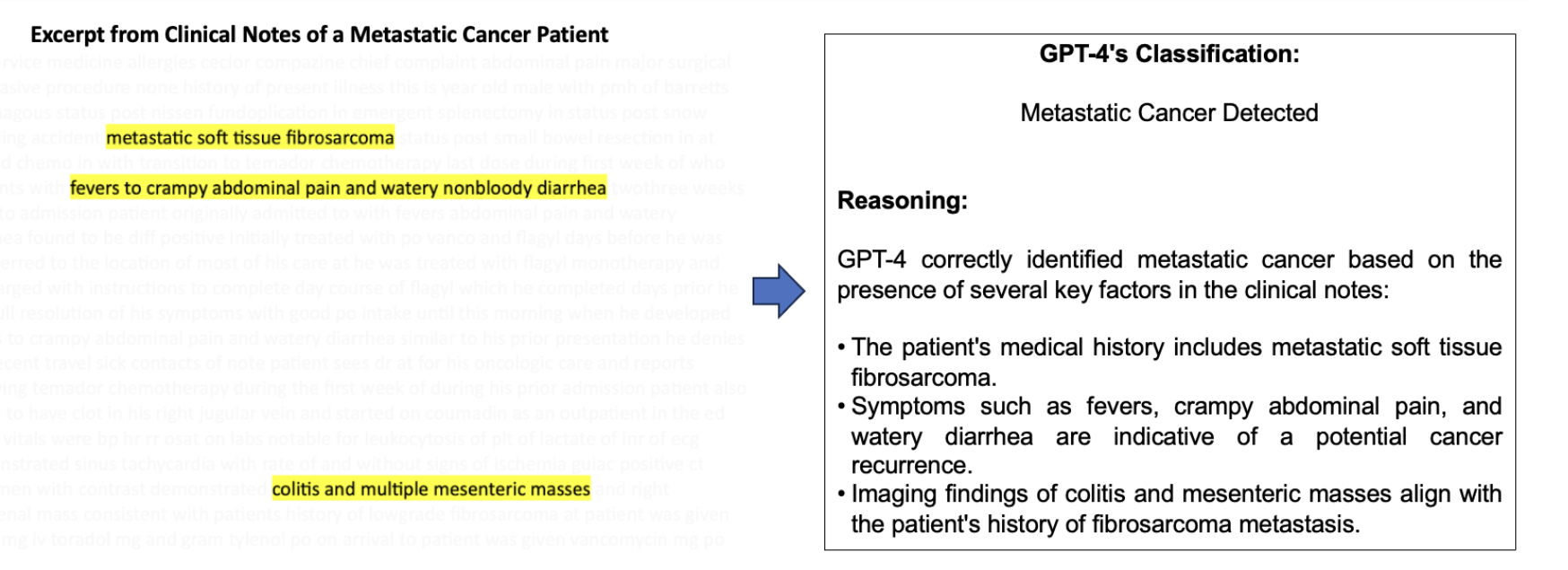
Example of Discharge Summaries for a Patient with Metastatic Cancer.

Next, we used a single label/example for the one-shot approach. The prompt accompanying this label/example was similar to the prompt 4 in the zero-shot approach, as it yielded better results. An input size of 1500 tokens for each combination, resulted in an F1 score of 0.228 for GPT-3.5 Turbo and 0.824 for GPT-4.

### Llama Fine-Tuning

Next, we explored the fine-tuning in Llama-7B. The performance metrics for the Llama fine-tuning models, as outlined in **Table 3**, indicate a range of effectiveness across different prompts. Specifically, prompts 1, 3 and 4 show the highest F1 scores at 0.600, accompanied by a recall of 0.500 and the highest precision of 0.750. The worst performance among the Llama fine-tuning models is for prompt 0, which has the lowest F1 score of 0.400 and recall of 0.286. However, its precision is relatively higher at 0.667. This indicates that while prompt 0 is precise when it does identify relevant cases, it misses a significant number of relevant instances. All these results suggest that while some fine-tuning of Llama models yields promising results, particularly in precision, there is room for improvement, especially when compared to the in-context zero-shot learning capabilities of models like GPT-4 as well as the fine-tuned PubMedBERT. This points towards a potential avenue for enhancing the fine-tuning strategies for LLMs, especially considering the accessibility and cost-effectiveness of open-source models such as Llama for research purposes.

**Table 3:** Performance Metrics for Llama-7B Fine-tuning Models.

| Model | Prompt ID | F1 Score | Recall | Precision |
| --- | --- | --- | --- | --- |
| Llama-7B | Prompt 0 | 0.400 | 0.286 | 0.667 |
|  | Prompt 1 | **0.600** | 0.500 | 0.750 |
|  | Prompt 2 | 0.400 | 0.333 | 0.500 |
|  | Prompt 3 | **0.600** | 0.500 | 0.750 |
|  | Prompt 4 | **0.600** | 0.500 | 0.750 |
|  | Prompt 5 | 0.500 | 0.429 | 0.600 |

### Impact of Key Parameters on GPT Performance

In our analysis of GPT-4’s reasoning abilities under various conditions, we utilized prompt 4 across a set of 100 test samples. We explored the impact of temperature settings, ranging from 0.1 to 1, on the performance of the model. Interestingly, we did not observe any variation in performance as we varied the temperature settings. This stability suggests that the reasoning task required for metastatic cancer identification is robust to such parameter adjustments.

The significance of keywords is well-established in the context of both machine learning models and human decision-making. To test the robustness of LLMs, we selectively removed key terms such as “metasta”, “cancer”, “advanced” and “metastatic” from the input text. The term “metasta” covers variations like metastasize, metastasis, and metastatic. Remarkably, GPT-4’s reasoning performance when guided by prompt 4 displayed remarkable resilience. We found that the removal of individual keywords from the input did not significantly affect the model’s reasoning capabilities.

Encouraged by the observation, we further experimented by randomly removing a specific percentage of tokens from the input text, while maintaining the original sequence of tokens. Our experiments revealed that the model’s performance remained stable, even with the removal of up to 40% of the tokens. These findings are in alignment with our earlier findings, where a reduction in token counts showed a better performance in the comparisons between inputs of differing token lengths (**Table 2**). Such resilience to information sparsity has added a compelling dimension to our understanding of LLM robustness in real-world scenarios, where incomplete or sparse data is a common challenge.

**Figure 2** illustrates the correlation between the degree of sparsity in the input text and the model’s performance. For each sparsity level tested, we replicated the experiment five times to ensure statistical reliability and added error bars to the resulting average F1 scores, thereby providing a clearer representation of the variability in our results.

**Figure 2.**
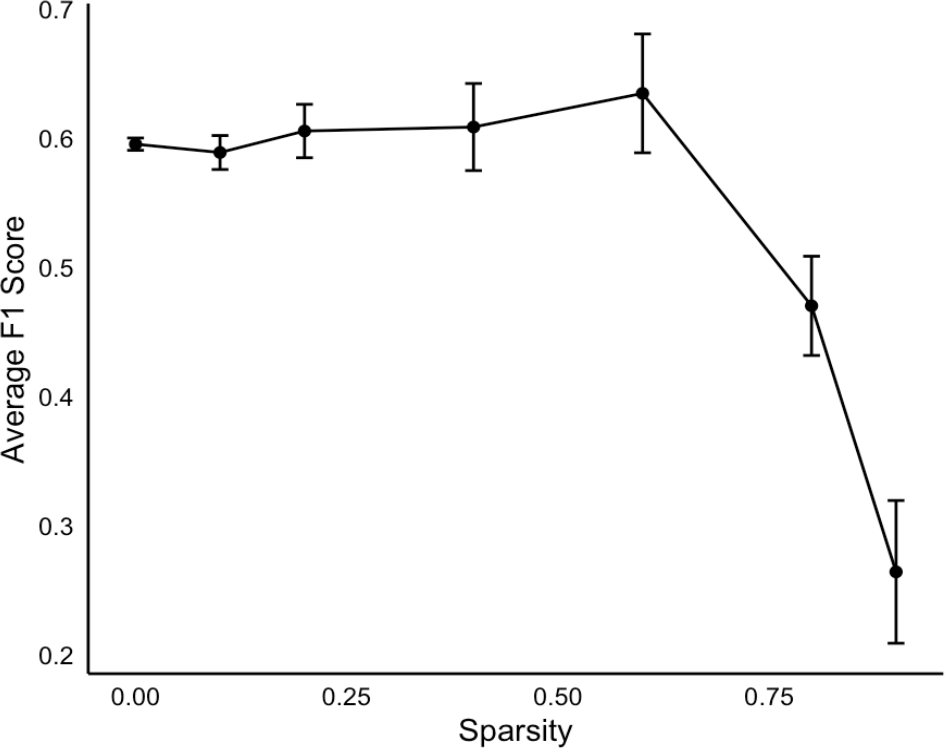
Correlation with Sparsity and Model Performance.

## Discussion

Through an in-depth study of one common task, we have demonstrated the enormous potential of applying LLMs in real clinical settings, owing to their remarkable performance and effectiveness. At the same time, we recognize the need for human expert intervention in certain challenging cases. After reviewing several false-negative instances identified by medical fellows, we suspect that evidence of metastatic cancer is absent in the discharge summaries. If such cases were removed from the test set, we expect that the overall performance of both LLMs and human annotators would improve. We did not observe hallucinations likely because of the nature of this reasoning task.

We found that a well-crafted prompt in zero-shot learning can yield performance that is comparable to, or even better than, that achieved through one-shot learning and fine-tuning strategies in this task. The addition of “*Let’s think step by step*” couldn’t compete in comparison to simple and clear prompts. The performance of the models (GPT-3.5 Turbo, GPT-4 and Llama-7b) correlates with the number of parameters in the pre-trained models. Although we did not investigate Llama-2, which has 65 billion parameters, we anticipate that significant effort is needed to optimize the fine-tune process. The Google BARD^29^ model was also tested using internal data, but provided no comparable results, so we proceeded without further investigation. Remarkably, the PubMedBERT model did not maintain its superior performance over the GPT-4 model. This observation suggests that the powerful abilities of GPT-4, when effectively harnessed through advanced prompt engineering strategies, can outperform specialized models that have undergone extensive domain-specific fine-tuning. This is potentially due to their large scale, diverse training data, and advanced architectures. This highlights the potential of leveraging the broad foundational knowledge encoded in LLMs to achieve remarkable accuracy in highly specialized fields like biomedicine, without the necessity for model specialization.

We observed that using the first 1,500 tokens yielded better results in most cases than using the first 3,000 tokens in the GPT-3.5Turbo model. This is likely because the essential information for diagnosis, such as patient medical history, is generally found at the beginning of the note, while the latter part often contains irrelevant information like medication history. This suggests that segmenting relevant sections of the notes or using the summarized text from the models like BART could not only reduce costs but also enhance performance. Additionally, we found that removing a significant number of tokens or keywords related to metastatic cancer did not adversely affect performance. This finding is particularly important given that our input text is often incomplete. It also implies that we could achieve comparable performance using a smaller subset of tokens. GPT-4 demonstrates superior performance in processing longer input sequences, maintaining high precision and recall, as evidenced by its optimal handling of 3000-token inputs across various prompts. Lastly, we noted that by instructing the model to provide concise answers either “Yes” or “No” at the end of the prompt can avoid lengthy responses thus facilitating downstream processing. Overall, this study provides practical guidelines for the use of LMM in the biomedical field.

## Data Availability

The MIMIC III database used in this study is publicly available online upon request.

https://physionet.org/content/mimiciii/1.4/

## Author contributions

B.C. conceived and supervised the study, conducted the robustness test, and wrote the manuscript. X.Z. coordinated the research, and the manual annotations. X.Z. and N.T. implemented the GPT models. S.V. prepared datasets. S.A. and J.W. implemented fine-tuning in Llama, H.M. conducted explorations on Google Bard, S.M. implemented PubMedBERT, D.L., A.D., and D.J. participated in manual annotation. M.W. helped prepare datasets. D.C. and J.Z. provided resources. All authors participated in manuscript preparation.

