## Supplemental Appendix A for "Comparison of Prompt Engineering and Fine-Tuning Strategies in Large Language Models in the Classification of Clinical Notes"

### Supplemental File

#### Appendix A: Cohort Construction for Advanced/Metastatic Cancer Data Set

This appendix details the cohort construction methodology utilized in our study, with a focus on Advanced/Metastatic Cancer within the MIMIC-III database. The approach follows the established protocol as described in the work[23].

The MIMIC-III database includes de-identified clinical data from over 53,000 hospital admissions in the intensive care units (ICU) of the Beth Israel Deaconess Medical Center between 2001 and 2012. Among the various types of clinical notes available in MIMIC-III, discharge summaries were chosen for their comprehensive representation of patient phenotypes. Specifically, discharge summaries from the first ICU visit of 'frequent flyer' patients, defined as those with three or more ICU admissions within a 365-day period, were extracted. Additionally, subsequent summaries from these patients and random summaries from non-frequent flyers were included.

The selected notes underwent annotation for ten specific phenotypes and Advanced/Metastatic Cancer is one of the phenotypes. Each note received at least two annotations per phenotype to ensure accuracy. The annotating team comprised two clinical researchers, two junior medical residents, two senior medical residents, and one practicing intensive care medicine physician. In cases of uncertainty, the final label was determined by one of the senior clinicians. To guarantee the quality of the labels and minimize errors, a rigorous annotation process was followed, which included double labeling and senior clinician review where necessary.

The cohort constructions in our research are directly aligned with those in the original work, ensuring a robust foundation for our analysis and subsequent findings.
